# Rare loss-of-function variants in *POLD1, PMS1* and *FAN1* modify age at onset of motor symptoms in Huntington’s disease

**DOI:** 10.64898/2026.06.16.26354747

**Authors:** Sahar Gelfman, Rujin Wang, Adrian I. Campos, Jae Hoon Sul, Qingqin S. Li, Silvia Alvarez, Vijay Kumar Pounjara, Chen Wang, Talaha Ali, Yuxin Zou, Anthony Marcketta, Arkopravo Ghosh, Kyoko Watanabe, Alexander Lachmann, Keyrun Adhikari, Andrey Ziyatdinov, Sean Yu, Amelia Averitt, Ashley Paynter, Michelle LeBlanc, Marcus Jones, Regeneron Genetics Center, Jonathan Marchini, Gonçalo R. Abecasis, Luca A. Lotta, Aris Baras, Seung Kwak, Jim Rosinski, Thomas F. Vogt, Manuel A. R. Ferreira, Eli A. Stahl, Giovanni Coppola

## Abstract

Huntington’s disease is a rare neurodegenerative disease whose primary risk factors are inherited expansions of a CAG repeat tract in the *HTT* gene. Somatic expansion of these tracts leads to neuronal toxicity, neuronal death and clinical disease progression. To identify genetic factors with a major impact on disease onset and progression, we genome sequenced 18,825 individuals for the ENROLL-HD study. Our results show rare inactivating mutations in three genes, all involved in DNA damage repair, are major determinants of age of onset for motor symptoms (n=10,610) and other clinical manifestations. Heterozygote carriers of predicted loss-of-function (pLoF) variants in *POLD1* and *PMS1* developed motor symptoms an average 20 years (n=3; P=1×10^−5^) and 7 years (n=6; P=2×10^−3^) later than non-carriers, respectively. Conversely, heterozygote carriers of pLoF variants in *FAN1* (n=30) developed symptoms 10 years earlier (P=2×10^−10^). Our findings highlight therapeutic strategies and help predict age of onset for at-risk individuals.

## MAIN TEXT

HD is a neurodegenerative disease occurring in individuals who inherit a pathogenic (≥40) CAG repeats in exon 1 of the huntingtin gene (*HTT*). The age at which disease symptoms appear is strongly determined by the size of the CAG expansion, with longer tracts associated with earlier onset^1^. While HTT germline repeats may be used to predict age of onset, there is still considerable variability in age of onset among individuals with the same number of repeats, indicating the existence of modifying factors. Recent genome-wide association studies have shown associations with age of onset for common genetic variants near genes involved in DNA damage response (DDR) or mismatch repair (MMR) pathways^2-4^. These findings, together with the observation that inherited CAG repeats expand somatically over decades and eventually become toxic and cause neuronal death^5^, raise the intriguing possibility that modulating the function of DDR and MMR genes could alter the course of disease. However, it remains unclear which specific genes should be targeted or whether their function should be inhibited or activated.

To better understand HD pathophysiology and support the identification of therapeutic targets, we performed whole-genome sequencing of 18,825 individuals in the ENROLL-HD cohort^6^, creating a valuable resource for HD research. Here we describe the first set of analyses conducted using this resource. All the results reported in this paper are supported by two independent and parallel analyses of sequence data for the ENROLL-HD cohort, carried out by teams of investigators at the Regeneron Genetics Center and the CHDI foundation (**Supplementary Note**, Li et al. 2026 *MedRxiv*). For our main analyses, we focused on 10,610 individuals with pathogenic CAG repeats in *HTT* and documented age at motor symptoms onset. Genome-wide common-variant analyses identified associations at nine loci outside of the *HTT* locus (**Supplementary Figure 1**), all previously reported^4^ and including seven located near DNA mismatch repair genes: *PMS1, MLH1, MSH3, PMS2, MLH3, FAN1* and *LIG1* (**Supplementary Figure 2**).

We next analyzed rare coding variants, which typically have larger effect sizes and provide for unequivocal causal gene identification. We identified three genes outside of the *HTT* locus where rare coding variants are significantly associated with motor symptom onset (see **Methods** and **Supplementary Figure 3**), two delaying (*POLD1* and *PMS1*) and one hastening (*FAN1*) onset (**Figure 1**). The largest delaying effect on motor onset was observed for rare coding variants in *POLD1* (overall omnibus gene-based test P [or P_gene_] = 7.9×10^−10^). *POLD1* encodes the proofreading subunit of DNA polymerase-delta. Remarkably, heterozygote carriers of ultra-rare predicted loss-of-function (pLoF) variants in *POLD1* (n=3) developed motor symptoms on average 20 years later than non-carriers who inherited the same number of CAG repeats (age at motor symptoms increased by +2.10 standard deviation units [SD] for carriers of *POLD1* pLoF variants with frequency < 0.1%, 95% CI 1.17 to 3.03, P=9.6×10^−6^; **Figure 1**). Smaller but still highly significant delays in motor onset were observed for heterozygote carriers of missense variants in *POLD1:* 5.5 years for rare deleterious missense variants (n=11; +1.05 SD for carriers of missense with frequency < 0.1%, 95% CI 0.56 to 1.53, P=2.6×10^−5^), and 2.2 years for the common missense variant Arg30Trp (19:50398939:C:T, n=208; +0.30 SD, 95% CI 0.19 to 0.41, P=1.4×10^−7^; **Figure 1**). The Arg30Trp missense variant was recently associated with age at total functional capacity score equal to 6 (TFC6)^7^, but it was unclear until now whether Arg30Trp results in loss or gain of *POLD1* function. Our findings strongly suggest that Arg30Trp causes partial loss of *POLD1* function, as its direction of effect aligns with that of pLoF variants. Together, our findings for pLOF variants, rare missense variants and Arg30Trp further reveal a dose-response relationship between the predicted level of *POLD1* inactivity and delay in motor symptoms onset (**Supplementary Figure 4**). This genetic association is in line with the role of *POLD1* in strand-displacement synthesis during mismatch repair, which can lead to repeat expansion ^8,9^ (**Figure 2**).

**Figure 1.**
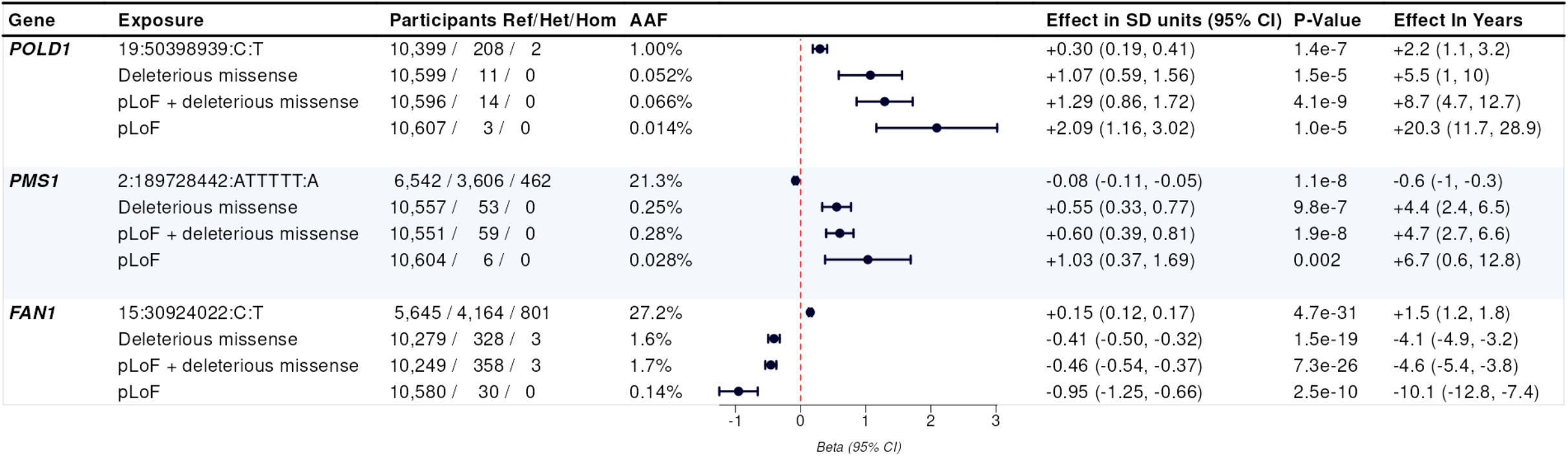
Association between age at onset of motor symptoms and genetic variants in *POLD1, PMS1* and *FAN1*. For each gene, we show genetic variants with the most significant association with age at onset of motor symptoms, considering (i) individual common variants included in the GWAS (first row); and (ii) rare coding variants tested on aggregate through gene-based tests (subsequent rows). Among the latter, the most significant association for each gene was observed when testing jointly pLoF and missense variants, specifically top 10% most deleterious missense for *POLD1* (with AAF<0.1%); top 20% most deleterious for *PMS1* (AAF<0.5%); and top 50% most deleterious for *FAN1* (AAF<1%). Effect in SD units was estimated using REGENIE after normalizing age at onset within groups of individuals with the same CAG repeat length, while effect in years was estimated from a linear model applied prior to normalization but including CAG repeat length as a categorical covariate.

**Figure 2.**
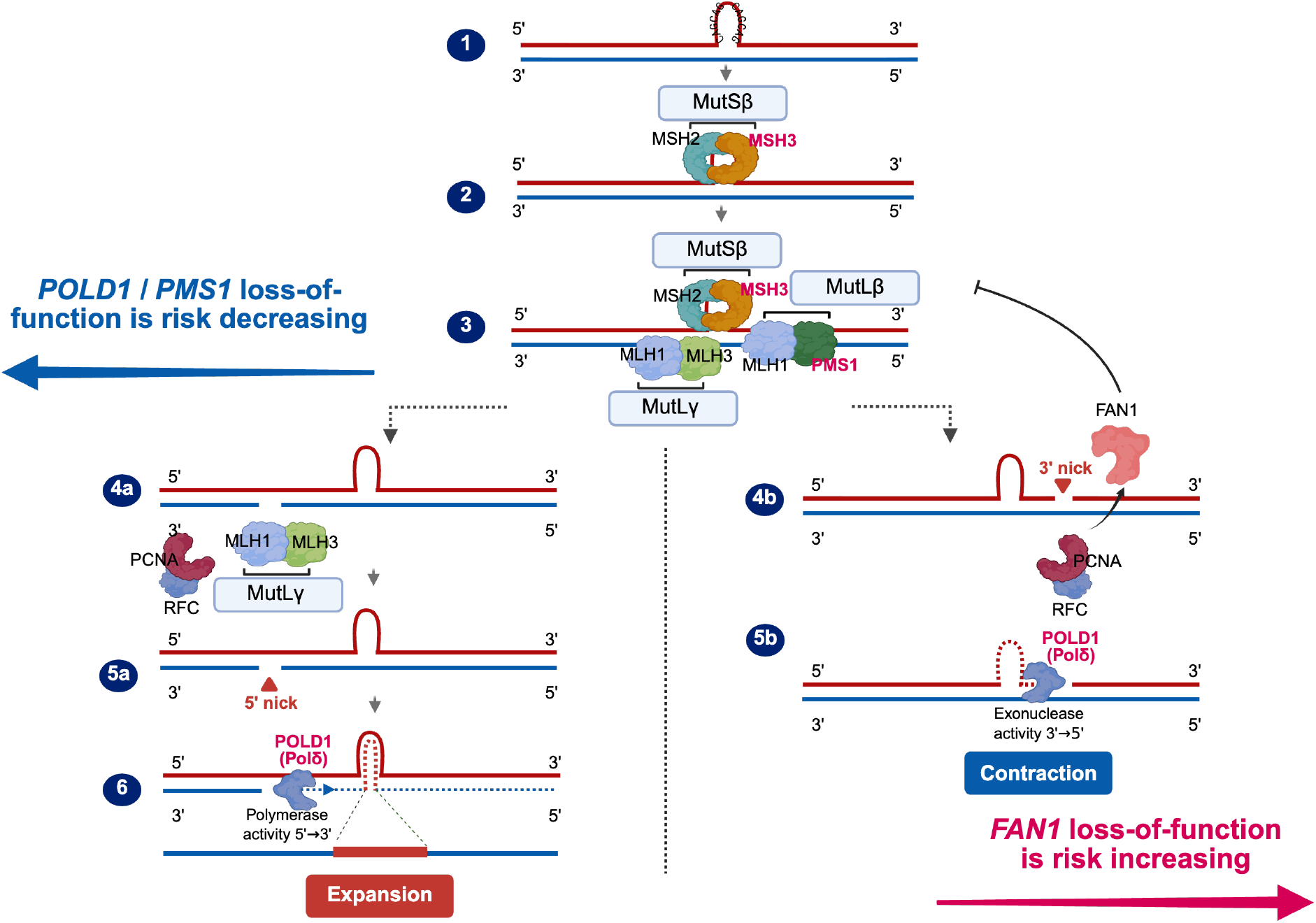
Inhibition of DNA mismatch repair genes may slow or prevent the expansion of toxic somatic CAG repeats in HD. 1) CAG/CTG loops are generated in the process of transcription and oxidative stress in postmitotic neurons^21^. **2)** CAG/CTG loops are preferentially recognized by MutSβ (MSH2/MSH3)^22,23^. **3)** MutSβ (MSH2/MSH3) recruits MutLγ (*MLH1/MLH3*) and MutLβ (*MLH1/PMS1*) - PMS1 might have a facilitating role potentially stabilizing MutSβ or the MutSβ–MutLγ complex^24,25^. **4a)** RFC-loaded PCNA regulates MutLγ endonuclease activity confining incisions near the loop 8-10^9,26^. **5a)** MutLγ endonuclease activity nicks the DNA opposite to the loop at the 5’ boundary ^26^ **6)** POLδ 5’→3’ strand displacement and synthesis toward loop includes the looped-out DNA, driving expansion^9,14,24^. **4b)** RFC-loaded PCNA stimulates and restricts *FAN1* nuclease activity, such that *FAN1* nicks the loop strand 3’ of the loop and sterically blocks MutSβ-MutLγ interaction 8-10^9^. **5b)** POLδ 3’→5’ exonuclease into looped-out DNA, driving excision and repeat contraction^9,14,26,27^.

The second gene with rare coding variants delaying motor symptoms was *PMS1* (P_gene_=2.6×10^−7^), encoding the yeast post meiotic segregation increased homolog 1 component of the MutLβ mismatch recognition complex. As with *POLD1*, the delay on motor onset was progressively larger as the predicted deleterious effect of *PMS1* coding variants increased (**Figure 1** and **Supplementary Figure 4**), reaching a 6.7-year delay among heterozygote carriers of pLoF variants (n=6; +1.08 SD for carriers of pLoF with frequency < 0.5%, 95% CI 0.42 to 1.74, P=0.001). These findings conclusively establish a role for *PMS1* in the pathophysiology of HD and demonstrate that loss of *PMS1* function delays the onset of motor symptoms. The molecular function of human *PMS1* is not fully understood. *PMS1* binds to *MLH1* forming the MutLβ heterodimer^10^, and may have a cooperative structural role with MutSβ and MutLγ to promote nicking the DNA strand opposite the mismatch loop, favoring repeat expansion (**Figure 2**)^11^.

The third gene significantly associated with age at motor symptoms onset was *FAN1* (P_gene_=6.7×10^−27^), encoding the DNA FANCD2 and FANCI associated nuclease 1, with rare coding variants greatly hastening onset. Specifically, heterozygote carriers of rare pLoF variants in *FAN1* (n=30) developed motor symptoms on average 10 years earlier than non-carriers (−0.95 SD for carriers of pLoF with frequency < 1%, 95% CI −1.24 to −0.65, P=3.8×10^−10^), while the most deleterious class of missense variants hastened onset by ~4 years (−0.41 SD for carriers of deleterious missense variants with frequency <1%, 95% CI −0.50 to −0.32, P=2.1×10^−19^; **Figure 1** and **Supplementary Figure 4**). These genetic associations strongly suggest that loss of *FAN1* activity promotes the somatic expansion of CAG repeats, which is consistent with its role in disrupting the physical interaction between MutSβ and MutLγ, required for MutLγ-driven repeat expansion^9,12^ (**Figure 2**).

Overall, across *POLD1, PMS1* and *FAN1*, 431 of 10,610 individuals (4%) were heterozygote carriers of rare pLoF or deleterious missense variants with a large effect on motor symptoms onset (**Figure 1**). For these individuals, age at onset could be assessed more accurately by considering rare deleterious coding variants in these genes in addition to their inherited CAG repeat length (**Figure 3**). Notably, large-effect variants in these three genes were significantly depleted (*POLD1*: OR=0.23, P=0.014; *PMS1*: OR=0.25, P=1.4×10^−5^) or enriched (*FAN1*: OR=2.52, P=2.8×10^−10^) among the 10,610 individuals with documented motor symptoms when compared to 3,883 individuals with pathogenic repeats in HTT that have not yet developed motor symptoms (**Supplementary Figure 5**), also sequenced as part of the ENROLL-HD cohort. These results provide independent support for a role of *POLD1, PMS1* and *FAN1* in HD pathophysiology.

**Figure 3.**
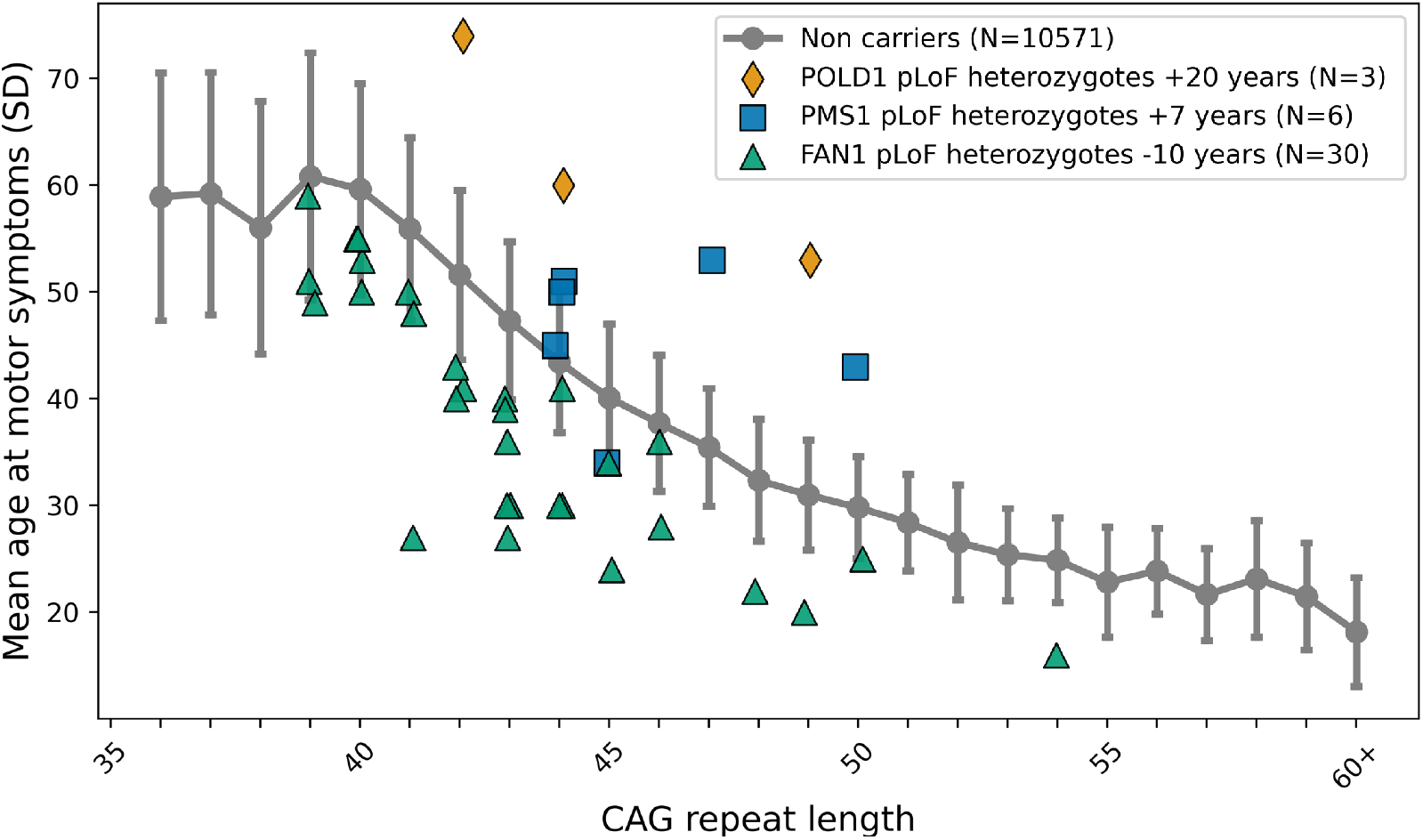
Age at motor symptom onset is greatly modified by pLoF variants in *POLD1, PMS1* and *FAN1*. Observed age at onset is shown for each individual carrier of a pLoF variant in the three genes, while the average age at onset (+/-standard deviation) is shown for non-carriers. Carrier datapoints with overlapping positions were randomly jittered along the x-axis.

Next, we tested if genetic variants in *POLD1, PMS1* and *FAN1* affected six clinical landmarks that occur throughout the disease course (**Supplementary Figure 6**). This includes the ages at Total Functional Capacity (TFC) 10 and 6 (TFC10, n=2,301; TFC6, n=1,702), and ages at HD integrated staging system 2 and 3 (HD-ISS2, n=2,469; HD-ISS3, n=9,380). Results were broadly consistent with the effects observed for the onset of motor symptoms: rare pLoFs in *POLD1* associated with an 18-year delay in TFC6 and 20-year delay in HD-ISS3 (**Supplementary Figure 7**), while rare pLoFs in *PMS1* delayed TFC6 by 6 years and HD-ISS3 by 8 years (**Supplementary Figure 8**). Carriers of rare pLoF variants in *FAN1* showed earlier onsets for nearly all clinical phenotypes tested (**Supplementary Figure 9**), including TFC6 (13 years earlier) and HD-ISS3 (9 years earlier). These results indicate that modulation of *POLD1, PMS1* and *FAN1* function also affects the age at which HD patients reach clinical landmarks later during disease course.

Thus far, we established that ~4% of HD cases carry rare deleterious coding variants in *POLD1, PMS1* or *FAN1* that have large effects on age at motor symptom onset, as well as later clinical landmarks. These results are consistent with HD mouse models wherein knocking out *PMS1* and *POLD1* prevents, while loss of *FAN1* promotes, somatic CAG repeat expansion^13,14^. Given these findings, and the observation that somatic expansion of CAG repeats leads to a low but steady rate of neuronal death^5^, we postulated that these three genes should also be associated with the rate of disease progression after onset. To test this, we quantified disease progression using longitudinal measures of the composite unified HD rating scale (cUHDRS), considering only symptomatic participants (with HD-ISS=2 or 3 and TFC<13; N=5,384), and tested this phenotype for association with *POLD1, PMS1* and *FAN1* variants (see **Methods**). For all three genes, the nearby common variants associated with age at onset of motor symptoms had nominally significant and directionally consistent effects on the rate of decline of cUHDRS (**Supplementary Figure 10)**. The largest effect was observed for Arg30Trp in *POLD1*, which slowed the rate of progression by 0.44 points (or about 39%) per year in heterozygote carriers (95% CI 0.26 to 0.61, P=1.6×10^−6^) when compared to non-carriers with the same number of CAG repeats. Rare coding variants in the three genes were not significantly associated with the rate of decline in cUHDRS, possibly due to low power. Similar results were observed with different clinical scores and analytical approaches (**Supplementary Note**). Taken together, these results support an effect of genetic variation in these genes on disease progression beyond motor onset.

In summary, we report large-effect genetic associations with HD onset in *POLD1, PMS1* and *FAN1*, which directly support the emerging model of HD pathophysiology whereby somatic *HTT* CAG repeat expansion throughout life can be hastened or slowed by changes in DDR and MMR function^5,13-15^ (**Figure 2**). Our results show not only that, among carriers of pathogenic *HTT* repeats, individuals with *FAN1* loss of function variants have much earlier onset than expected, those with *POLD1* or *PMS1* loss of function variants have later than expected disease onset – and in fact, are over-represented among individuals who have not yet manifested motor symptoms. These genetic associations thus identify actionable therapeutic targets that could help delay disease onset and help predict age at which motor symptoms are first expected.

## Methods

### Study populations

These studies were carried out in accordance with the ethical principles outlined in the Declaration of Helsinki, Good Clinical Practices guidelines, and applicable regulatory requirements. The study protocols were approved by the local, regional, or central Institutional Review Board (IRB) or Independent Ethics Committee (IEC) or Research Ethic Board (REB) overseeing the respective clinical sites. All participants provided written informed consent before enrollment. The data for analyzed subjects were provided by the participants in the Enroll-HD study and made available by the CHDI Foundation. Enroll-HD is a global clinical research platform designed to facilitate clinical research in Huntington’s Disease. We leveraged 18,825 participants from CHDI with available whole-genome sequencing and clinical phenotype data, including 1,019 non-European individuals.

### Clinical phenotypes

Our primary trait of interest was age at motor symptom onset, which was defined as the first time when motor symptoms manifested. We also examined age at HD diagnosis, age at TFC6, age at DCL4, and age at HD-ISS Stage 3. We defined additional landmarks based upon total motor score (TMS), symbol digit modalities test (SDMT), Stroop word reading test (SWRT), verbal fluency test (VERFCT), hospital anxiety and depression scale (HADS) and composite unified Huntington Disease rating scale (cUHDRS) scores. For all the age-related quantitative phenotypes, rank inverse-normal transformation (RINTing) was applied in each germline CAG repeat length specific subgroup. For the survival analysis of motor symptoms onset, participants with more than 35 CAG repeats and with no age at motor symptoms were included as censored at their maximum observed age. Our primary progression analysis looked at the change in cUHDRS in a subset of observations on symptomatic participants, and additional analyses are described in **Supplementary Note**. Briefly, we calculated individual progression rate as the rate per day between cUHDRS values at the first and last visits, after excluding asymptomatic visits where TFC score was 13 or ISS was 0 or 1. Progression rates were adjusted for baseline cUHDRS and age at motor symptoms onset, and the residuals were RINTed, within strata defined by CAG repeat, sex, and education (low = high school or less, high = more than high school).

### Whole genome sequencing, read mapping and variant calling

Whole genome sequencing (WGS) was performed at the Regeneron Genetics Center. DNA was extracted from lymphoblastoid cell lines (LCLs) and/or whole blood samples from participating cohorts. A high-throughput automated approach developed at Regeneron was used to prepare high-quality genomic DNA for whole-genome sequencing (WGS). Sequencing was performed using paired-end 150 bp reads on Illumina NovaSeq X platform. Sequencing had a mean coverage depth of 37.5X and 99.6% of samples were above 27X.

Sequencing reads from the whole genome assays in FASTQ format were generated from Illumina image data using bcl2fastq program (Illumina). Following the OQFE (original quality functional equivalent) protocol (Krasheninina et al., 2020), sequence reads were mapped to GRCh38 references using BWA MEM (Li et al., 2009) in an alt-aware manner, read duplicates were marked, and additional per-read tags were added. Single nucleotide variations (SNV) and short insertion and deletions (indels) were identified using a Parabricks accelerated version of DeepVariant v1.5 with the default WGS model and reported in per-sample genome VCF (gVCF). These exome gVCFs were aggregated with GLnext v0.12.2 using the pre-configured DeepVariantWGS setting (personal communications) into joint-genotyped multi-sample project-level VCF (pVCF), which was converted to bed/bim/fam format using PLINK 1.9 (Chang et al., 2015). We determined continental ancestry super-groups (African (AFR), admixed population (AMR), East Asian (EAS), European (EUR) and South Asian (SAS)) by projecting each sample onto reference principal components calculated from the HapMap3 reference panel^16^.

### Statistical Analyses

We leveraged whole-genome sequencing of more than 18,000 individuals from the ENROLL-HD cohort, to conduct GWAS of various clinical landmark phenotypes and discover novel genetic modifiers of Huntington’s Disease. Association analyses were performed using mixed-effects linear regression models (for quantitative traits), or Firth bias-corrected logistic or cox regression models (for binary traits and survival analyses, respectively) implemented in REGENIE^17^. Analyses were adjusted for age, sex, age-squared, age-by-sex and age-squared-by-sex interaction terms, the first 10 common-variant-derived principal components, the first 20 rare-variant-derived principal components, and a polygenic score which robustly adjusted for relatedness and population structure approximated using REGENIE. To ensure independence of signals generated by common and rare variants, we performed conditional analyses with adjustment for common variants.

Gene-based tests of rare coding variants were also performed using REGENIE. Briefly, for a given gene, we tested each phenotype for association with non-synonymous variants having at least one carrier and a frequency <1%. A total of 49 different types of gene-based tests were performed per gene-phenotype pair (**Supplementary Table 1**); for example, burden tests of variants grouped by their effect on gene function (e.g. pLoF variants, deleterious missense variants) and by their allele frequency (e.g. with a frequency <0.1%). The following variants were classified as pLoF: frameshift-causing indels, variants affecting splice acceptor and donor sites, and variants leading to stop gain. The deleterious effect of missense variants on gene function was predicted based on five large language models, as described below. Results from the 49 tests were then summarized into a single gene-level P-value as described previously^18,19^, which was used to identify significant associations between a gene and a phenotype (significance threshold set at P<0.05/19,478 genes tested=2.6×10^−6^).

### Predicting the deleterious effect of missense variants on gene function

To classify missense variants as benign or pathogenic, we integrated pathogenic scores obtained from five different large protein language models (LLMs), specifically scores from (i) three published LLMs, ESM-1v, ESM-1b, and AlphaMissense; and (ii) our refinement of predictions from ESM-1v and ESM-1b using proteomics data on 1,175 proteins across 46,665 individuals, as described previously^20^. To combine scores across these five LLMs, we did the following. First, the pathogenic score produced by each LLM was converted to a percentile score (i.e. ranked from 0 to 1) across all possible missense variants. Second, for each variant, the five percentile scores were averaged across the five LLMs. Third, the average percentile score was re-ranked across all missense variants, giving each variant a score from 0 (predicted to be most benign) to 1 (predicted to be most deleterious). This final prediction score was then used to define nested percentile-based thresholds, referred to as missense_50 (top 50%), missense_60 (top 40%), missense_70 (top 30%), missense_80 (top 20%), and missense_90 (top 10%), which progressively include missense variants with a higher probability of being deleterious. For example, missense_90 represents the group of missense variants with a prediction score among the top 10% across all missense variants.

## Supporting information

RGC banner

## Data Availability

All WGS data used in the present study are available upon request to the CHDI management

https://enroll-hd.org/for-researchers/data-from-human-samples/

## Code Availability

BWA-MEM used for sequence alignment is available as an open-source software at https://github.com/lh3/bwa. DeepVariant used for variant calling is available as an open-source software at https://github.com/google/deepvariant. PLINK 1.9 that was used for genotypic analysis is available at https://github.com/chrchang/plink-ng. The REGENIE software for whole genome regression that was used to perform all genetic association analysis is available at https://github.com/rgcgithub/regenie. R Statistical Computing 4.x and its libraries broom (1.0.7), data.table (1.17.0), dplyr (1.1.4), forestplot (3.1.3), reshape2 (1.4.4), ggplot2 (3.5.1), sqldf (0.4-11), stringi (1.8.4), stringr (1.5.1) and tidyverse (2.0) and Python 3.11.9 along with modules matplotlib (3.10.6), seaborn (0.13.2), statsmodels (0.14.2) and numpy (1.26.4) were used for data wrangling, visualization, and statistical analyses.

## Data Availability

Both whole genome sequencing data and phenotypic data used in this study is available from http://www.enroll-hd.org/. A data use agreement is needed to obtain the data used in this study.

