## Supplementary figures for "Rare loss-of-function variants in *POLD1, PMS1* and *FAN1* modify age at onset of motor symptoms in Huntington’s disease"

Supplementary Figure 1

A Summary results of GWAS of age at onset of motor symptoms among 10,610 individuals with pathogenic CAG repeats in *HTT*

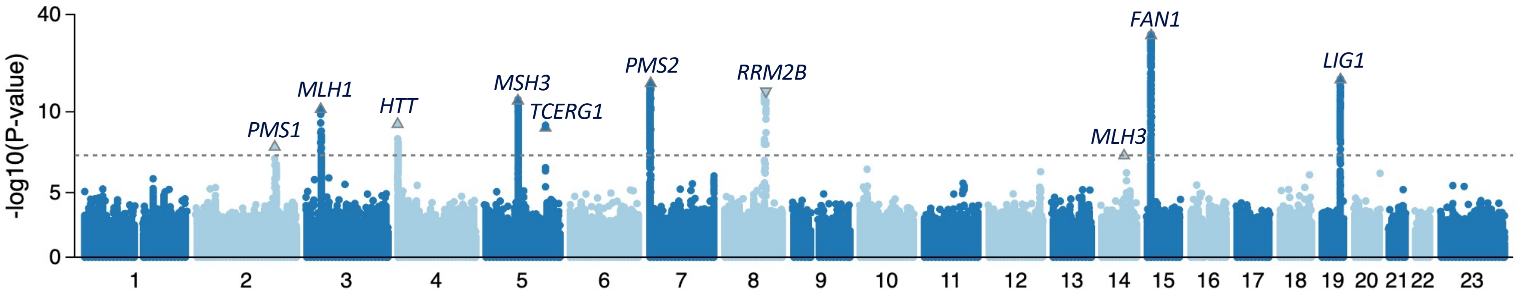


B Association between age at onset of motor symptoms and lead variant in ten loci identified in GWAS of AMO

| Gene | Name | Participants Ref/Het/Hom | MAF | Effect (95% CI) | P-Value | Years Het | Years Hom |
|---|---|---|---|---|---|---|---|
| RRM2B | 8:102204458:G:A | 9,018 / 1,530 / 62 | 7.8% | -0.16 (-0.20, -0.12) | 7.6e-14 | -1.08 | -3.60 |
| PMS1 | 2:189704465:C:A* | 6,542 / 3,452 / 505 | 21.2% | -0.08 (-0.11, -0.05) | 1.2e-8 | -0.50 | -1.15 |
| MLH3 | 14:75086187:AAAAAAT:A* | 2,682 / 5,241 / 2,663 | 49.9% | -0.06 (-0.08, -0.04) | 4.2e-8 | -0.48 | -0.72 |
| MSH3 | 5:80638411:G:T | 4,208 / 4,872 / 1,529 | 37.4% | +0.08 (0.06, 0.11) | 1.1e-12 | 0.69 | 1.53 |
| MLH1 | 3:37389270:A:G | 5,672 / 4,155 / 782 | 27.0% | +0.08 (0.06, 0.11) | 2.6e-11 | 0.65 | 1.13 |
| PMS2 | 7:5982995:C:T | 7,750 / 2,628 / 227 | 14.5% | +0.13 (0.10, 0.16) | 4.7e-16 | 0.91 | 2.32 |
| LIG1 | 19:48177968:G:A | 8,107 / 2,329 / 174 | 12.6% | +0.14 (0.11, 0.18) | 6.8e-17 | 0.91 | 2.48 |
| FAN1 | 15:30924022:C:T | 5,645 / 4,164 / 801 | 27.2% | +0.15 (0.12, 0.17) | 4.7e-31 | 1.52 | 2.28 |
| TCERG1 | 5:146507273:G:A | 10,068 / 535 / 7 | 2.6% | +0.22 (0.15, 0.29) | 8.0e-10 | 1.57 | 8.31 |
| HTT | 4:3099058:TATC:T | 10,243 / 364 / 2 | 1.7% | +0.27 (0.19, 0.36) | 4.7e-10 | 1.92 | 7.09 |

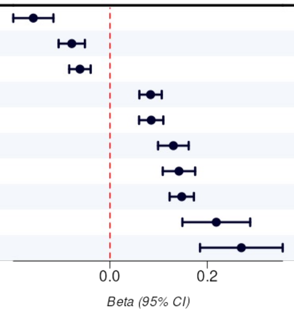


*: alt allele is the major allele

**Supplementary Figure 1.** Summary of association results from GWAS of age at onset of motor symptoms among individuals (*N*=10,610) with pathogenic *HTT* CAG repeats in the CHDI cohort. **A)** Manhattan plot showing association (-$\log_{10}$*P*-value) with common variants (MAF≥1%). The dotted grey line demarcates the genome-wide significance threshold of $P$=5x$10^{-8}$. **B)** Associations between age at onset of motor symptoms and lead variants in 10 loci identified from Figure 1A. Higher effect estimates correspond to a delayed age at onset. Effect in years are estimated for heterozygote and homozygote carriers, respectively.

Supplementary Figure 2

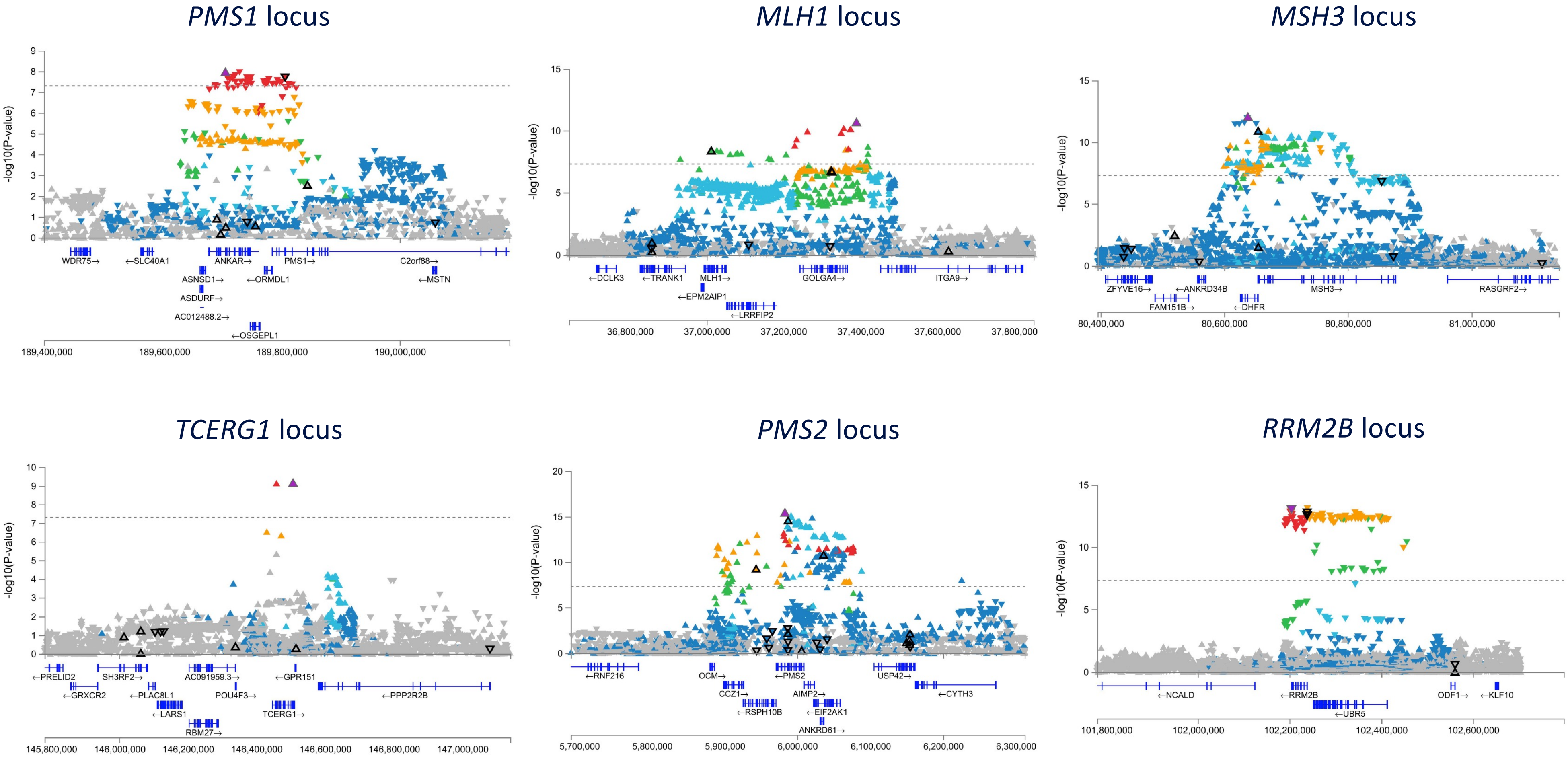

Supplementary Figure 2 (cont)

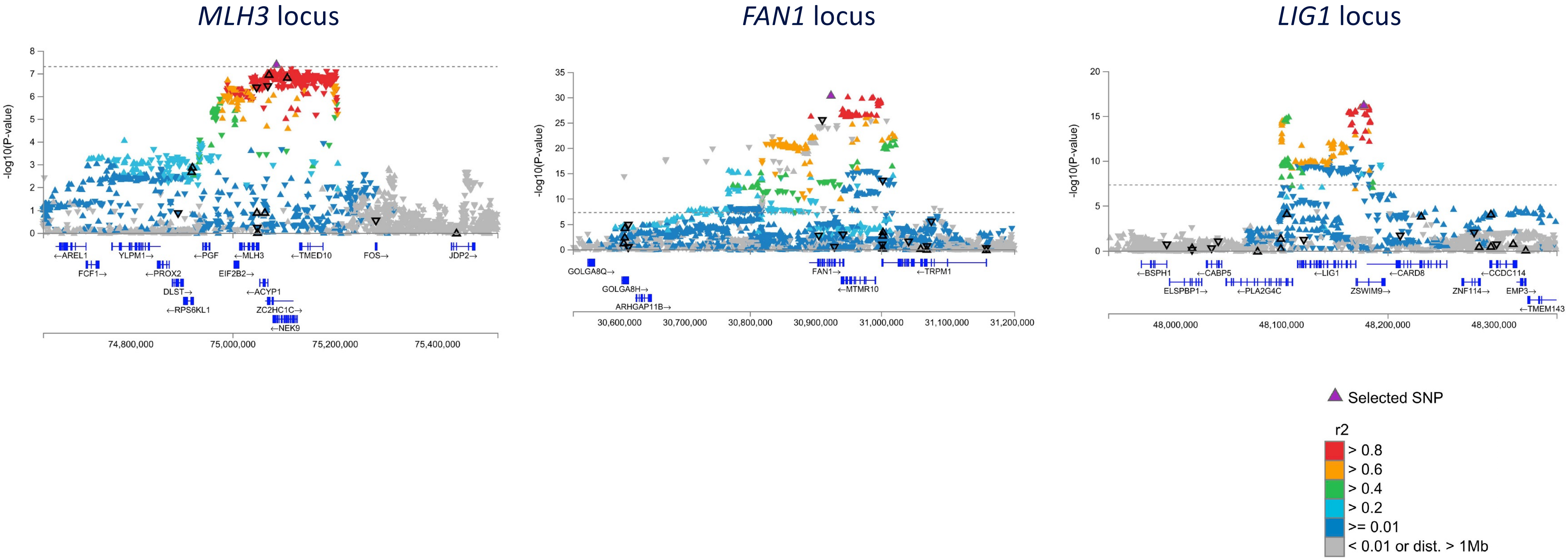


**Supplementary Figure 2.** Regional associations plots for the genome-wide significant loci from GWAS of age at onset of motor symptoms. Variants are colored based on their linkage disequilibrium with the lead variant (purple triangle). Upward facing triangles represent variants with effect size > 0, and downward facing triangles represent effect size < 0. Unadjusted *P*-values derived from Firth-regression (two-sided test) implemented in REGENIE.

Supplementary Figure 3

**Association between rare coding variants in 19,462 genes and age at onset of motor symptoms among 10,610 individuals with pathogenic CAG repeats in *HTT***

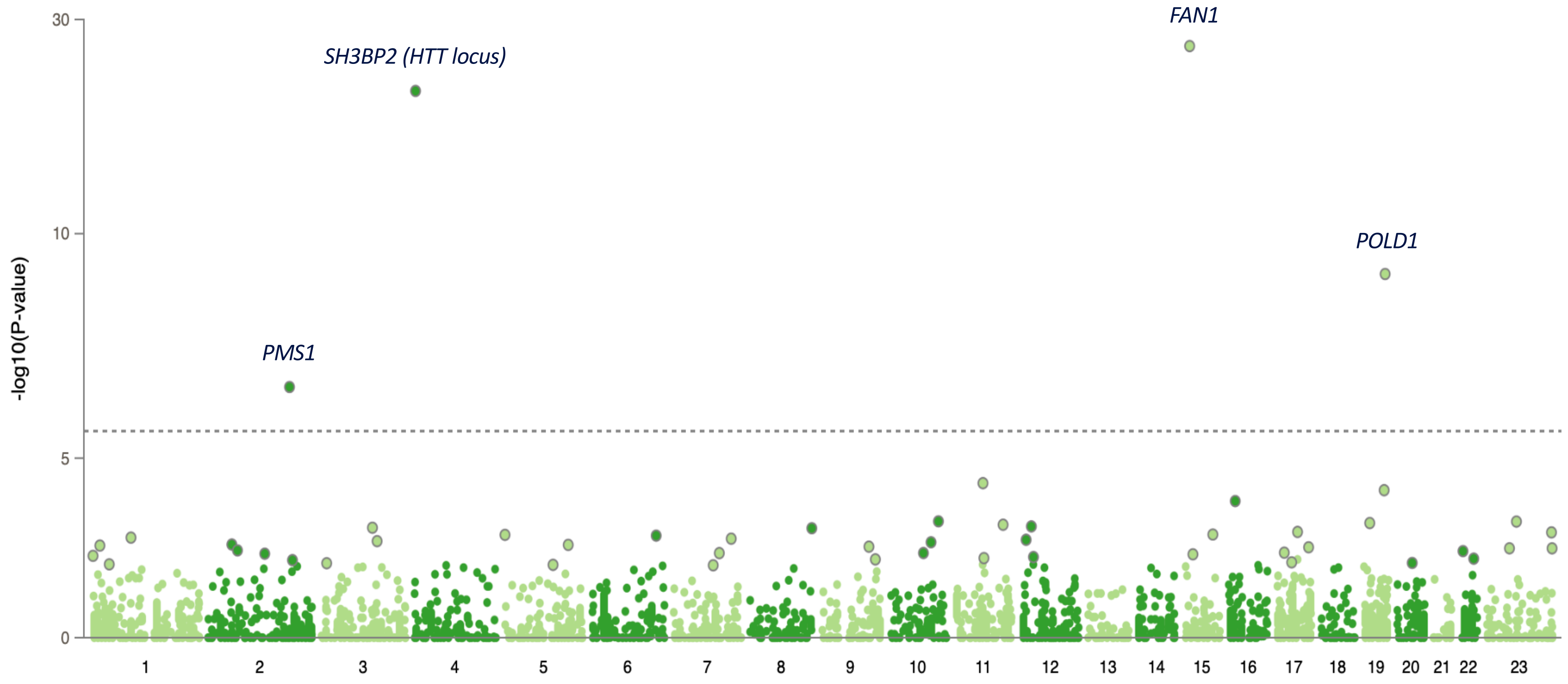


**Supplementary Figure 3.** Summary of association results between age at onset of motor symptoms and rare coding variants from whole genome sequencing in the CHDI cohort. We tested 26 million rare (AAF<1%) variants derived from whole genome sequencing of 10,610 individuals with pathogenic CAG repeats in HTT. Manhattan plots of rare coding variants tested on aggregate through an omnibus gene-based test (each point represents an overall omnibus gene-based test P for a gene; Methods).

### Supplementary Figure 4

| Gene | Exposure | Participants Ref/Het/Hom | AAF | Effect in SD units (95% CI) | P-Value | Effect In Years |
|---|---|---|---|---|---|---|
| ***POLD1*** | missense (>60%, <=70%), AAF<0.1% | 10,565 / 45 / 0 | 0.21% | -0.06 (-0.30, 0.18) | 0.620 | n.s. |
| | missense (>50%, <=60%), AAF<0.1% | 10,550 / 60 / 0 | 0.28% | -0.04 (-0.25, 0.17) | 0.711 | n.s. |
| | missense (>70%, <=80%), AAF<0.1% | 10,563 / 47 / 0 | 0.22% | +0.02 (-0.21, 0.26) | 0.847 | n.s. |
| | 19:50398939:C:T (rs3218772) p.Arg30Trp | 10,399 / 208 / 2 | 1.00% | +0.30 (0.19, 0.41) | 1.4e-7 | +2.2 (1.1, 3.2) |
| | missense (>80%, <=90%), AAF<0.1% | 10,593 / 17 / 0 | 0.080% | +0.48 (0.09, 0.87) | 0.016 | +2.4 (-1.3, 6) |
| | missense (>90%), AAF<0.1% | 10,599 / 11 / 0 | 0.052% | +1.07 (0.59, 1.56) | 1.5e-5 | +5.5 (1, 10) |
| | pLoF, AAF<0.1% | 10,607 / 3 / 0 | 0.014% | +2.09 (1.16, 3.02) | 1.0e-5 | +20.3 (11.7, 28.9) |
| ***PMS1*** | 2:189728442:ATTTTT:A (rs3067424) intronic | 6,542 / 3,606 / 462 | 21.3% | -0.08 (-0.11, -0.05) | 1.1e-8 | -0.6 (-1, -0.3) |
| | missense (>50%, <=60%), AAF<0.5% | 10,529 / 81 / 0 | 0.38% | -0.07 (-0.25, 0.11) | 0.419 | n.s. |
| | missense (>70%, <=80%), AAF<0.5% | 10,596 / 14 / 0 | 0.066% | +0.13 (-0.30, 0.56) | 0.567 | n.s. |
| | missense (>60%, <=70%), AAF<0.5% | 10,601 / 9 / 0 | 0.042% | +0.30 (-0.23, 0.84) | 0.268 | n.s. |
| | missense (>80%, <=90%), AAF<0.5% | 10,586 / 24 / 0 | 0.11% | +0.47 (0.14, 0.80) | 0.005 | +3.3 (0.2, 6.3) |
| | missense (>90%), AAF<0.5% | 10,581 / 29 / 0 | 0.14% | +0.62 (0.32, 0.92) | 5.1e-5 | +5.4 (2.6, 8.2) |
| | pLoF, AAF<0.5% | 10,604 / 6 / 0 | 0.028% | +1.03 (0.37, 1.69) | 0.002 | +6.7 (0.6, 12.8) |
| ***FAN1*** | missense (>70%, <=80%), AAF<1% | 10,360 / 247 / 3 | 1.2% | -0.32 (-0.42, -0.22) | 4.7e-10 | -3.1 (-4, -2.1) |
| | missense (>60%, <=70%), AAF<1% | 10,585 / 25 / 0 | 0.12% | -0.35 (-0.67, -0.03) | 0.033 | -4.5 (-7.5, -1.5) |
| | missense (>50%, <=60%), AAF<1% | 10,582 / 28 / 0 | 0.13% | -0.45 (-0.75, -0.14) | 0.004 | -4.2 (-7, -1.4) |
| | 15:30910758:G:A (rs150393409) p.Arg507His | 10,354 / 255 / 0 | 1.2% | -0.55 (-0.66, -0.45) | 4.4e-26 | -4.7 (-5.7, -3.8) |
| | missense (>80%, <=90%), AAF<1% | 10,577 / 33 / 0 | 0.16% | -0.56 (-0.85, -0.28) | 8.7e-5 | -6.7 (-9.3, -4.1) |
| | missense (>90%), AAF<1% | 10,589 / 21 / 0 | 0.099% | -0.80 (-1.15, -0.45) | 8.8e-6 | -7.1 (-10.3, -3.8) |
| | pLoF, AAF<1% | 10,580 / 30 / 0 | 0.14% | -0.95 (-1.25, -0.66) | 2.5e-10 | -10.1 (-12.8, -7.4) |

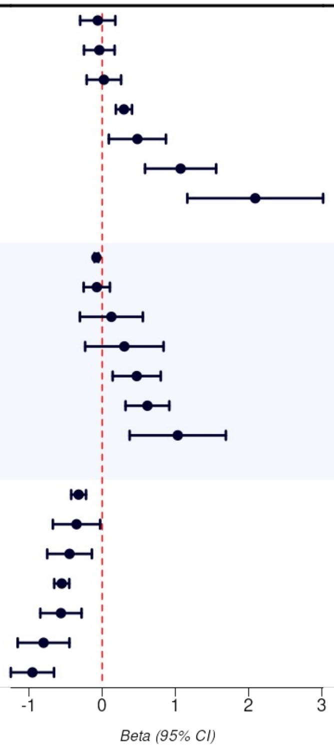


**Supplementary Figure 4.** Forest plot of associations between genetic variation within or near the three genes identified by gene-based tests and age at motor symptom onset of Huntington's disease. For each gene we show the strongest missense variant in the locus (if not present, we show the strongest common variant signal), the burden masks with increasingly non-cumulative predicted-deleterious missense rare variants, and the burden mask of putative loss of function (pLoF) rare variants; sorted by effect sizes. Effect in years and their confidence intervals are estimated from a linear model including CAG repeats as a categorical covariate.

Supplementary Figure 5

| Gene | Exposure | Cases Ref/Het/Hom | Controls Ref/Het/Hom | AAF | OR (95% CI) | P-Value |
|---|---|---|---|---|---|---|
| ***POLD1*** | 19:50398939:C:T | 10,399 / 208 / 2 | 3,773 / 108 / 2 | 1.1% | 0.67 (0.47, 0.94) | 0.022 |
| | deleterious missense | 10,599 / 11 / 0 | 3,876 / 7 / 0 | 0.062% | 0.50 (0.11, 2.19) | 0.358 |
| | pLoF + deleterious missense | 10,596 / 14 / 0 | 3,873 / 10 / 0 | 0.083% | 0.23 (0.07, 0.74) | 0.014 |
| | pLoF | 10,607 / 3 / 0 | 3,880 / 3 / 0 | 0.021% | 0.02 (0.00, 0.21) | 0.001 |
| ***PMS1*** | 2:189728442:ATTTTT:A | 6,542 / 3,606 / 462 | 2,482 / 1,244 / 157 | 21.0% | 1.18 (1.08, 1.29) | 4.5e-4 |
| | deleterious missense | 10,557 / 53 / 0 | 3,848 / 35 / 0 | 0.30% | 0.28 (0.15, 0.53) | 1.0e-4 |
| | pLoF + deleterious missense | 10,551 / 59 / 0 | 3,841 / 42 / 0 | 0.35% | 0.25 (0.13, 0.47) | 1.4e-5 |
| | pLoF | 10,604 / 6 / 0 | 3,875 / 8 / 0 | 0.048% | 0.06 (0.01, 0.39) | 0.003 |
| ***FAN1*** | 15:30924022:C:T | 5,645 / 4,164 / 801 | 1,950 / 1,590 / 343 | 27.7% | 0.76 (0.70, 0.82) | 5.9e-11 |
| | deleterious missense | 10,279 / 328 / 3 | 3,779 / 103 / 1 | 1.5% | 2.15 (1.60, 2.89) | 4.5e-7 |
| | pLoF + deleterious missense | 10,249 / 358 / 3 | 3,775 / 107 / 1 | 1.6% | 2.52 (1.89, 3.36) | 2.8e-10 |
| | pLoF | 10,580 / 30 / 0 | 3,879 / 4 / 0 | 0.12% | 26.51 (7.66, 91.80) | 2.3e-7 |

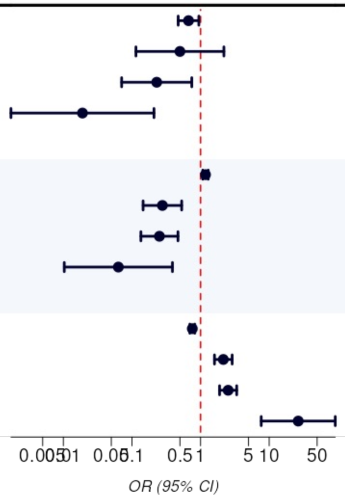


**Supplementary Figure 5.** Forest plot of associations between genetic variation within or near the three genes identified by gene-based tests and a case-control analysis of developing motor symptoms ever (10,610 cases and 3,883 controls). For each gene we show the same variant and mask displayed in Figure 1. The deleterious missense burden masks are missense (>90%), AAF<0.1% for POLD1; missense (>80%), AAF<0.5% for PMS1; missense (>50%), AAF<1% for FAN1.

Supplementary Figure 6

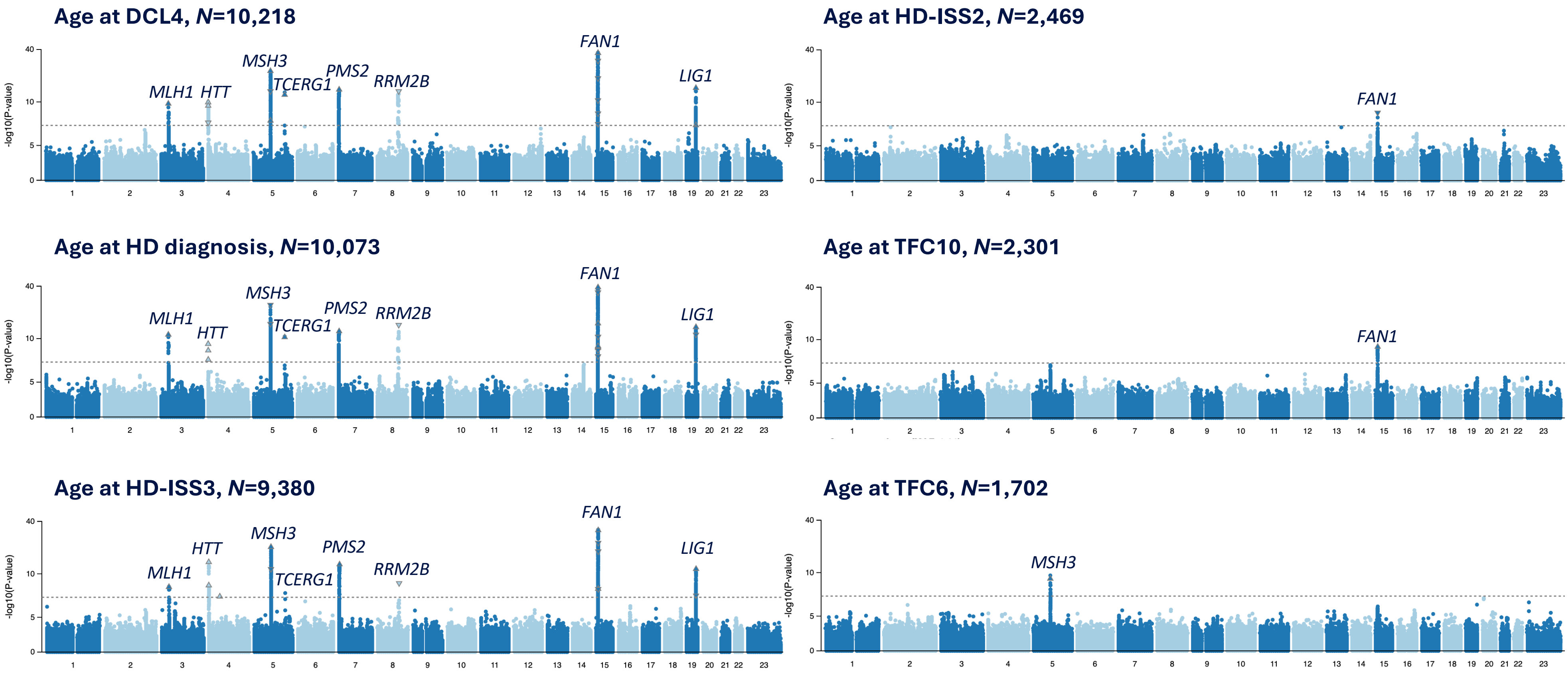


**Supplementary Figure 6.** Summary of association results from GWASs of other clinical landmarks among individuals with pathogenic *HTT* CAG repeats in the CHDI cohort. Manhattan plot showing association (-$\log_{10}$*P*-value) with common variants (MAF≥1%). The dotted grey line demarcates the genome-wide significance threshold of *P*=5x$10^{-8}$.

Supplementary Figure 7

### POLD1

| Pheno | Participants Ref/Het/Hom | AAF | Effect in SD units (95% CI) | P-Value | Het Effect In Years |
|---|---|---|---|---|---|
| ***19:50398939:C:T*** | | | | | |
| age at dcl 4 | 10,020 / 195 / 2 | 0.97% | +0.21 (0.11, 0.31) | 6.5e-5 | +2.2 (1, 3.3) |
| age at hd diagnosis | 9,883 / 187 / 2 | 0.95% | +0.34 (0.23, 0.45) | 4.3e-9 | +3.3 (2.2, 4.4) |
| age at hdiss stage 2 | 2,413 / 55 / 0 | 1.1% | +0.03 (-0.15, 0.20) | 0.772 | n.s. |
| age at hdiss stage 3 | 9,196 / 182 / 1 | 0.98% | +0.24 (0.14, 0.33) | 1.6e-6 | +2.1 (0.9, 3.3) |
| age at motor symptoms | 10,399 / 208 / 2 | 1.00% | +0.30 (0.19, 0.41) | 1.4e-7 | +2.2 (1.1, 3.2) |
| age at tfcscore 10 | 2,259 / 41 / 0 | 0.89% | +0.29 (0.07, 0.50) | 0.010 | +3.4 (1.1, 5.7) |
| age at tfcscore 6 | 1,678 / 24 / 0 | 0.71% | +0.51 (0.20, 0.81) | 0.001 | +4.3 (1.5, 7) |
| ***Deleterious missense*** | | | | | |
| age at dcl 4 | 10,208 / 10 / 0 | 0.049% | +0.75 (0.30, 1.21) | 0.001 | +4.3 (-0.6, 9.2) |
| age at hd diagnosis | 10,064 / 9 / 0 | 0.045% | +1.05 (0.53, 1.57) | 8.6e-5 | +6.5 (1.4, 11.6) |
| age at hdiss stage 2 | 2,468 / 1 / 0 | 0.020% | +2.01 (0.74, 3.27) | 0.002 | +13.8 (-3.1, 30.6) |
| age at hdiss stage 3 | 9,368 / 12 / 0 | 0.064% | +0.54 (0.16, 0.92) | 0.005 | +2 (-2.7, 6.7) |
| age at motor symptoms | 10,599 / 11 / 0 | 0.052% | +1.07 (0.59, 1.56) | 1.5e-5 | +5.5 (1, 10) |
| age at tfcscore 10 | 2,299 / 2 / 0 | 0.043% | +1.16 (0.19, 2.13) | 0.019 | +9.7 (-0.8, 20.1) |
| age at tfcscore 6 | 1,700 / 2 / 0 | 0.059% | +1.62 (0.58, 2.66) | 0.002 | +16.8 (7.4, 26.2) |
| ***pLoF*** | | | | | |
| age at dcl 4 | 10,215 / 3 / 0 | 0.015% | +1.63 (0.80, 2.46) | 1.2e-4 | +20.2 (11.3, 29.1) |
| age at hd diagnosis | 10,071 / 2 / 0 | 0.010% | +1.71 (0.60, 2.82) | 0.002 | +18.1 (7.3, 28.9) |
| age at hdiss stage 3 | 9,377 / 3 / 0 | 0.016% | +1.52 (0.76, 2.27) | 8.4e-5 | +20.3 (10.9, 29.6) |
| age at motor symptoms | 10,607 / 3 / 0 | 0.014% | +2.09 (1.16, 3.02) | 1.0e-5 | +20.3 (11.7, 28.9) |
| age at tfcscore 6 | 1,701 / 1 / 0 | 0.029% | +1.71 (0.24, 3.18) | 0.023 | +17.9 (4.6, 31.2) |

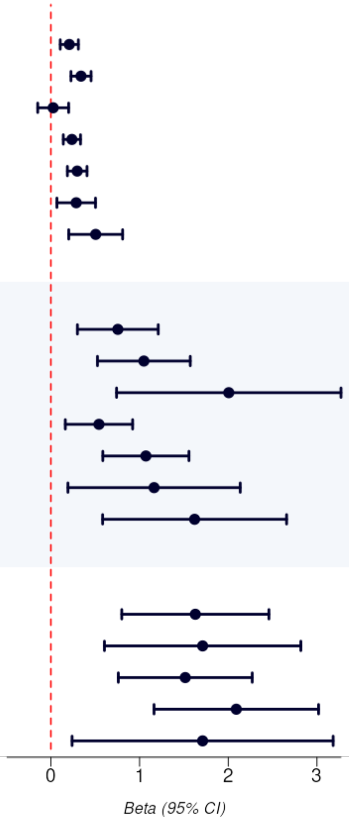


**Supplementary Figure 7. Forest plot of associations between common and rare variants in *POLD1* and clinical landmarks of Huntington's disease. Top panel**) The associations of common variants near POLD1 and clinical landmarks show a consistent effect in early and later stage clinical landmarks. **Middle panel**) The associations of burden masks consisting of POLD1 deleterious missense rare variants exhibit larger effects than common variants. **Lower panel**) The associations of burden masks consisting of POLD1 pLoF variants exhibit larger effects with less power than deleterious missense variants.

Supplementary Figure 8

### PMS1

| Pheno | Participants Ref/Het/Hom | AAF | Effect in SD units (95% CI) | P-Value | Het Effect In Years |
|---|---|---|---|---|---|
| ***2:189728442:ATTTTT:A*** | | | | | |
| age at dcl 4 | 6,306 / 3,476 / 436 | 21.3% | -0.06 (-0.09, -0.04) | 3.6e-7 | -0.6 (-1, -0.3) |
| age at hd diagnosis | 6,217 / 3,417 / 439 | 21.3% | -0.07 (-0.10, -0.04) | 6.3e-7 | -0.4 (-0.8, -0.1) |
| age at hdiss stage 2 | 1,545 / 817 / 107 | 20.9% | -0.05 (-0.09, -0.00) | 0.030 | -0.4 (-1.1, 0.4) |
| age at hdiss stage 3 | 5,780 / 3,195 / 405 | 21.3% | -0.06 (-0.08, -0.03) | 3.6e-6 | -0.7 (-1, -0.3) |
| age at motor symptoms | 6,542 / 3,606 / 462 | 21.3% | -0.08 (-0.11, -0.05) | 1.1e-8 | -0.6 (-0.9, -0.3) |
| age at tfcscore 10 | 1,422 / 781 / 98 | 21.2% | -0.07 (-0.12, -0.02) | 0.008 | -0.6 (-1.2, 0.1) |
| age at tfcscore 6 | 1,053 / 576 / 73 | 21.2% | -0.10 (-0.17, -0.04) | 0.001 | -0.8 (-1.5, -0.1) |
| ***Deleterious missense*** | | | | | |
| age at dcl 4 | 10,168 / 50 / 0 | 0.24% | +0.42 (0.21, 0.62) | 5.9e-5 | +4.6 (2.4, 6.8) |
| age at hd diagnosis | 10,021 / 52 / 0 | 0.26% | +0.49 (0.28, 0.71) | 8.9e-6 | +4.2 (2.1, 6.3) |
| age at hdiss stage 2 | 2,451 / 18 / 0 | 0.36% | +0.25 (-0.06, 0.56) | 0.118 | n.s. |
| age at hdiss stage 3 | 9,331 / 49 / 0 | 0.26% | +0.41 (0.22, 0.59) | 2.1e-5 | +4.2 (1.8, 6.5) |
| age at motor symptoms | 10,557 / 53 / 0 | 0.25% | +0.55 (0.33, 0.77) | 9.8e-7 | +4.5 (2.4, 6.5) |
| age at tfcscore 10 | 2,291 / 10 / 0 | 0.22% | -0.08 (-0.51, 0.36) | 0.726 | n.s. |
| age at tfcscore 6 | 1,692 / 10 / 0 | 0.29% | +0.63 (0.16, 1.10) | 0.009 | +5.3 (1, 9.5) |
| ***pLoF*** | | | | | |
| age at dcl 4 | 10,211 / 7 / 0 | 0.034% | +0.96 (0.42, 1.51) | 5.2e-4 | +7.4 (1.5, 13.2) |
| age at hd diagnosis | 10,066 / 7 / 0 | 0.035% | +1.23 (0.64, 1.82) | 4.7e-5 | +6.3 (0.5, 12.1) |
| age at hdiss stage 2 | 2,467 / 2 / 0 | 0.041% | +0.03 (-0.86, 0.93) | 0.941 | n.s. |
| age at hdiss stage 3 | 9,374 / 6 / 0 | 0.032% | +0.94 (0.40, 1.48) | 5.9e-4 | +8.2 (1.6, 14.9) |
| age at motor symptoms | 10,604 / 6 / 0 | 0.028% | +1.03 (0.37, 1.69) | 0.002 | +6.7 (0.6, 12.8) |
| age at tfcscore 6 | 1,701 / 1 / 0 | 0.029% | +0.75 (-0.72, 2.23) | 0.317 | n.s. |

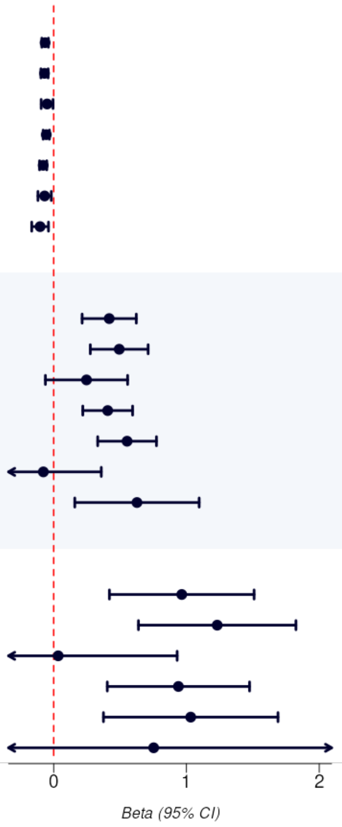


**Supplementary Figure 8**. **Forest plot of associations between common and rare variants in *PMS1* and clinical landmarks of Huntington's disease. Top panel**) The associations of common variants near PMS1 and clinical landmarks show a consistent effect in early and later stage clinical landmarks. **Middle panel**) The associations of burden masks consisting of PMS1 deleterious missense rare variants exhibit larger effects than common variants. **Lower panel**) The associations of burden masks consisting of PMS1 pLoF variants exhibit larger effects with less power than deleterious missense variants.

Supplementary Figure 9

**FAN1**

| Pheno | Participants Ref/Het/Hom | AAF | Effect in SD units (95% CI) | P-Value | Het Effect In Years |
|---|---|---|---|---|---|
| ***15:30924022:C:T*** | | | | | |
| age at dcl 4 | 5,431 / 4,005 / 782 | 27.3% | +0.14 (0.12, 0.17) | 2.9e-35 | +1.6 (1.3, 1.9) |
| age at hd diagnosis | 5,352 / 3,950 / 771 | 27.3% | +0.17 (0.14, 0.19) | 1.2e-39 | +1.5 (1.2, 1.9) |
| age at hdiss stage 2 | 1,327 / 948 / 194 | 27.1% | +0.11 (0.06, 0.15) | 3.5e-7 | +1.5 (0.8, 2.2) |
| age at hdiss stage 3 | 4,975 / 3,686 / 719 | 27.3% | +0.13 (0.11, 0.15) | 1.0e-30 | +1.7 (1.3, 2) |
| age at motor symptoms | 5,645 / 4,164 / 801 | 27.2% | +0.15 (0.12, 0.17) | 4.7e-31 | +1.5 (1.2, 1.8) |
| age at tfcscore 10 | 1,216 / 918 / 167 | 27.2% | +0.14 (0.10, 0.19) | 1.2e-9 | +1.4 (0.7, 2) |
| age at tfcscore 6 | 892 / 693 / 117 | 27.2% | +0.13 (0.07, 0.19) | 7.0e-6 | +1.6 (0.9, 2.3) |
| ***Deleterious missense*** | | | | | |
| age at dcl 4 | 9,896 / 319 / 3 | 1.6% | -0.38 (-0.46, -0.30) | 5.9e-20 | -4.1 (-4.9, -3.2) |
| age at hd diagnosis | 9,755 / 315 / 3 | 1.6% | -0.40 (-0.48, -0.31) | 2.3e-18 | -4.1 (-5, -3.2) |
| age at hdiss stage 2 | 2,402 / 66 / 1 | 1.4% | -0.31 (-0.47, -0.16) | 6.6e-5 | -3.3 (-5.4, -1.2) |
| age at hdiss stage 3 | 9,105 / 273 / 2 | 1.5% | -0.29 (-0.37, -0.21) | 1.2e-12 | -3.6 (-4.6, -2.6) |
| age at motor symptoms | 10,279 / 328 / 3 | 1.6% | -0.41 (-0.50, -0.32) | 1.5e-19 | -4.1 (-4.9, -3.2) |
| age at tfcscore 10 | 2,237 / 64 / 0 | 1.4% | -0.31 (-0.49, -0.14) | 4.2e-4 | -3.1 (-5, -1.2) |
| age at tfcscore 6 | 1,649 / 53 / 0 | 1.6% | -0.69 (-0.90, -0.48) | 5.4e-11 | -6.1 (-8, -4.3) |
| ***pLoF*** | | | | | |
| age at dcl 4 | 10,190 / 28 / 0 | 0.14% | -0.83 (-1.11, -0.56) | 2.7e-9 | -10.9 (-13.8, -7.9) |
| age at hd diagnosis | 10,046 / 27 / 0 | 0.13% | -0.77 (-1.08, -0.47) | 6.6e-7 | -9 (-12, -6.1) |
| age at hdiss stage 2 | 2,462 / 7 / 0 | 0.14% | -0.29 (-0.77, 0.19) | 0.240 | n.s. |
| age at hdiss stage 3 | 9,356 / 24 / 0 | 0.13% | -0.63 (-0.90, -0.36) | 4.9e-6 | -11 (-14.3, -7.7) |
| age at motor symptoms | 10,580 / 30 / 0 | 0.14% | -0.95 (-1.25, -0.66) | 2.5e-10 | -10.1 (-12.8, -7.4) |
| age at tfcscore 10 | 2,299 / 2 / 0 | 0.043% | -0.24 (-1.21, 0.73) | 0.623 | n.s. |
| age at tfcscore 6 | 1,696 / 6 / 0 | 0.18% | -0.92 (-1.52, -0.31) | 0.003 | -13.5 (-18.9, -8) |

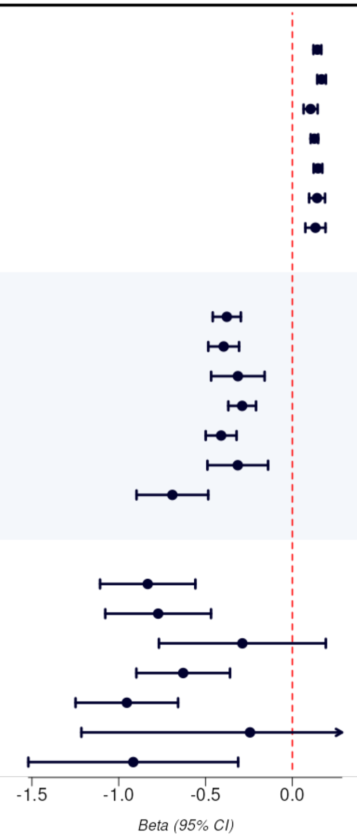


**Supplementary Figure 9**. **Forest plot of associations between common and rare variants in *FAN1* and clinical landmarks of Huntington's disease. Top panel**) The associations of common variants near FAN1 and clinical landmarks show a consistent effect in early and later stage clinical landmarks. **Middle panel**) The associations of burden masks consisting of FAN1 deleterious missense rare variants exhibit larger effects than common variants. **Lower panel**) The associations of burden masks consisting of FAN1 pLoF variants exhibit larger effects with less power than deleterious missense variants.

Supplementary Figure 10

| Gene | Exposure | Participants Ref/Het/Hom | AAF | Effect (95% CI) | P-Value |
|---|---|---|---|---|---|
| ***POLD1*** | 19:50398939:C:T | 5,272 / 111 / 0 | 1.0% | +0.44 (0.26, 0.61) | 1.6e-6 |
| | deleterious missense | 5,377 / 7 / 0 | 0.065% | +0.30 (-0.40, 1.01) | 0.394 |
| | pLoF + deleterious missense | 5,376 / 8 / 0 | 0.074% | +0.34 (-0.32, 0.99) | 0.314 |
| | pLoF | 5,380 / 1 / 0 | 0.009% | +0.56 (-1.29, 2.41) | 0.553 |
| ***PMS1*** | 2:189728442:ATTTTT:A | 3,329 / 1,836 / 219 | 21.1% | -0.07 (-0.11, -0.02) | 0.003 |
| | deleterious missense | 5,361 / 23 / 0 | 0.21% | +0.04 (-0.35, 0.43) | 0.834 |
| | pLoF + deleterious missense | 5,357 / 27 / 0 | 0.25% | +0.01 (-0.35, 0.37) | 0.950 |
| | pLoF | 5,380 / 4 / 0 | 0.037% | -0.16 (-1.09, 0.77) | 0.734 |
| ***FAN1*** | 15:30924022:C:T | 2,866 / 2,115 / 403 | 27.1% | +0.06 (0.02, 0.10) | 0.002 |
| | deleterious missense | 5,213 / 169 / 2 | 1.6% | -0.11 (-0.25, 0.03) | 0.135 |
| | pLoF + deleterious missense | 5,200 / 182 / 2 | 1.7% | -0.11 (-0.25, 0.03) | 0.114 |
| | pLoF | 5,371 / 13 / 0 | 0.12% | -0.13 (-0.65, 0.38) | 0.613 |

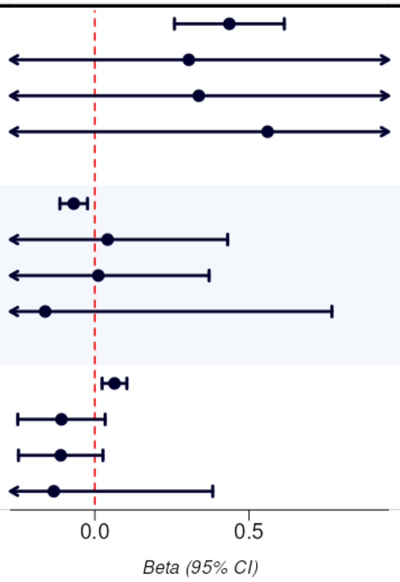


**Supplementary Figure 10.** Forest plot of genetic associations with cUHDRs progression restricting to symptomatic participants, excluding data points where TFC score is 13 or ISS is 0 or 1. Results are adjusted for baseline value and age at motor symptom, with the outcome first RINTed by CAG, sex and education.
