## Supplementary Information for "Rare loss-of-function variants in *POLD1, PMS1* and *FAN1* modify age at onset of motor symptoms in Huntington’s disease"

### Supplementary Note

#### **Estimating genetic effects on age at motor symptoms onset in years**

Association analyses of age at motor symptoms onset were performed using REGENIE (which accounts for population substructure and relatedness) after applying a rank-based inverse normal transformation in groups of individuals with the same CAG repeat length. This approach captures a substantial amount of variation in age at onset and therefore increases power for association testing. However, after applying the phenotype transformation within CAG repeat groups, the effect sizes estimated in genetic association analyses are expressed in standard deviation (SD) units and not in years (which is directly interpretable). To obtain a genetic effect expressed in years, we used two different approaches, which yielded very similar estimates:

- Approach 1. Within each CAG group:
  1. Estimate the mean age at onset for non-carriers.
  2. Calculate the difference between each carrier and the mean of non-carriers.
  3. Estimate the mean (and standard error) of the difference.
- Approach 2. Fit an ordinary least squares (OLS) model of the type y~g+X+e where y is age at motor symptom onset in years, g is the genetic instrument of interest and X is a dummy matrix specifying the sample belongs to a specific CAG repeat group. The effect sizes retain the raw scale and can be interpreted as the effect of carrier status while keeping CAG repeats constant.

Our empirical tests have yielded extremely concordant results between these two approaches (**Figure S1** below). Given the known behavior of a linear model, the ease of interpretation and the better handling of residual degrees of freedom, we have decided to report the OLS estimates and their confidence intervals. Nonetheless, it is very important to highlight that the utility of a standard error or confidence interval goes towards the statistical significance of an association. We consider the REGENIE-based standard errors and confidence intervals to be the most adequate for that.

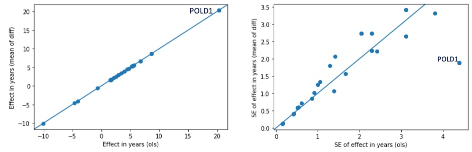

**Figure S1. Estimated effect sizes in years estimated with approach 1 (y-axis) and approach 2 (x-axis).** Results (effect on the left plot, standard error [SE] on the right plot) are shown for several burden tests. The diagonal line represents x=y.

#### **Disease progression traits**

Our main progression analysis looked at the change in the clinical variable cUHDRS in a subset of observations on symptomatic participants, but we also performed secondary analyses of progression for other clinical variables (see below). For a given clinical variable, we calculated the individual progression rate by taking the first visit value and the last visit value and computing the progression rate per day between these two visits. To ensure individuals experienced a symptom during progression, we excluded visits where TFC score is 13 or ISS is 0 or 1. Individuals are then stratified into each CAG repeat, sex, and education (low = high school or less, high = more than high school) group and their progression rates are corrected for baseline measurement and age-at-motor symptom. We performed rank-based inverse normal transformation on residuals of progression rates, which are then used in the genetic association analyses. Additional sensitivity analyses included (1) using this same strategy across other clinical variables including total motor score (TMS) and Total Functional Capacity score (TFC) (see **Figure S2**, below); and (2) testing for progression using the interaction between genotype and time since baseline in a linear mixed model, accounting for random intercepts and slopes per participant (see **Figure S3**, further below).

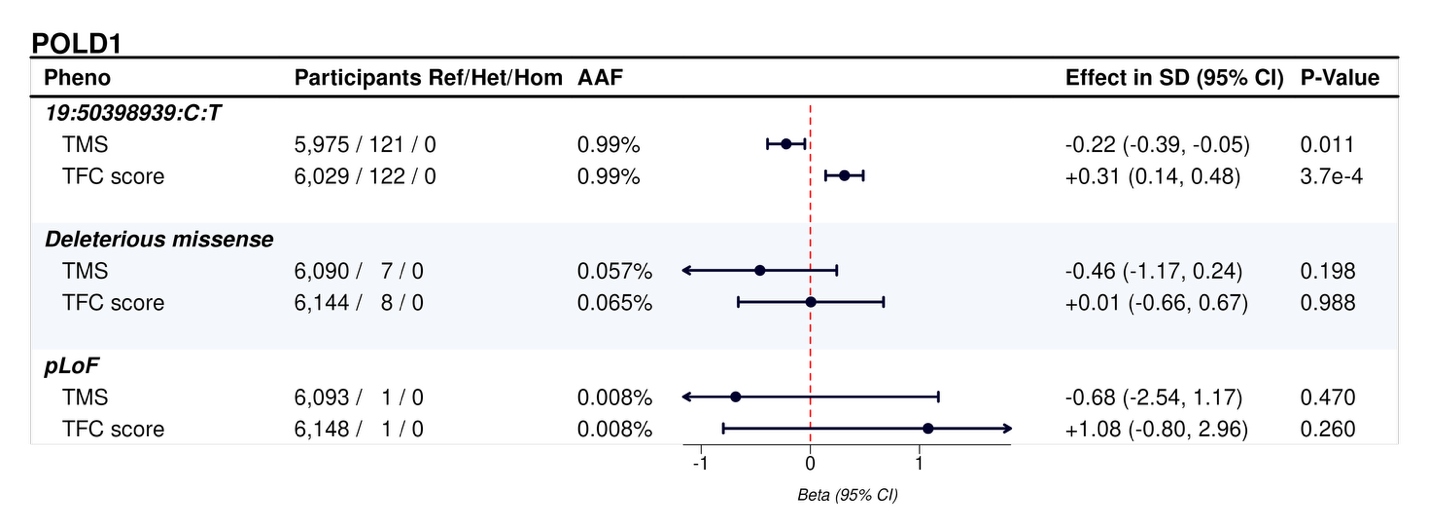

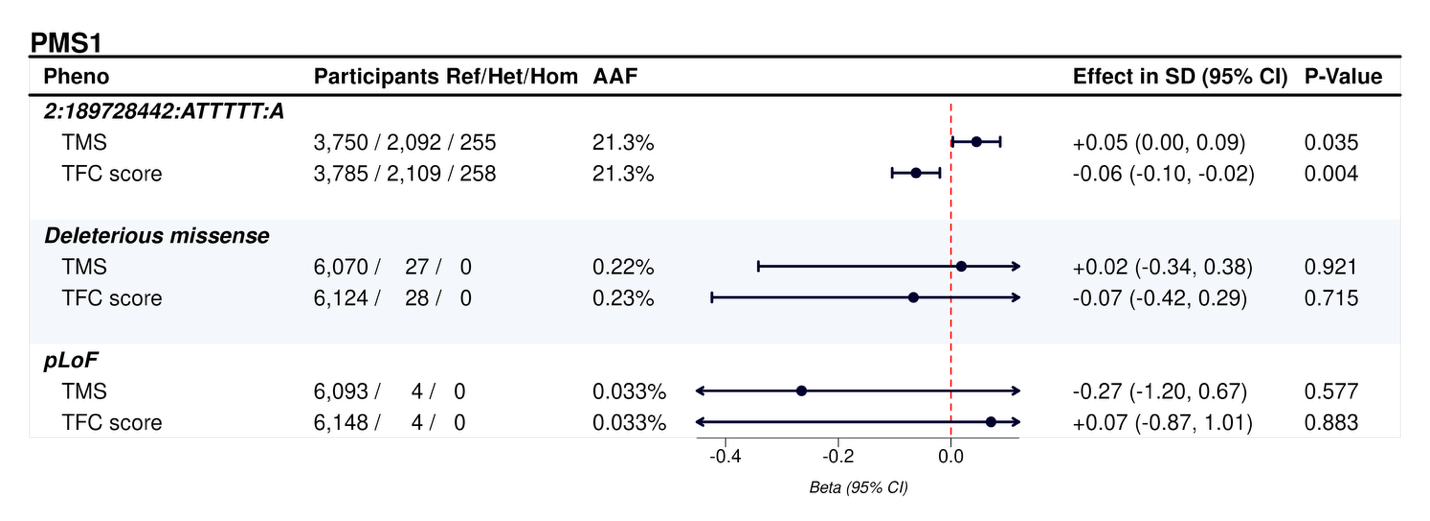

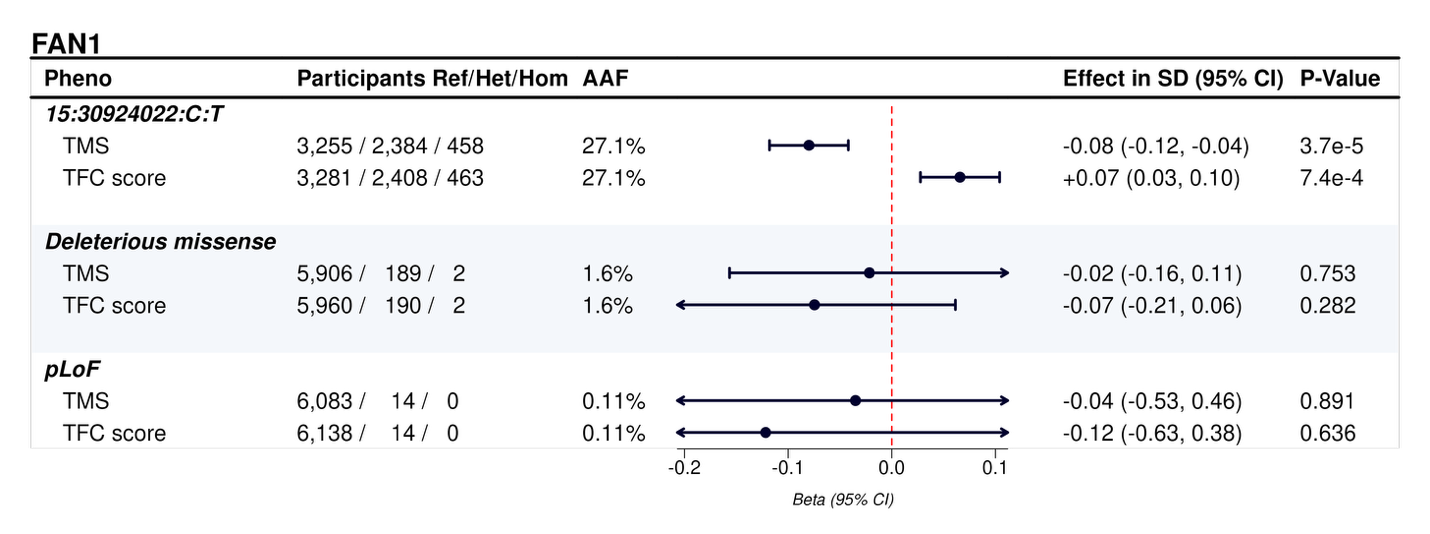

**Figure S2. Progression of other clinical traits.** Forest plot of genetic associations with progression of other clinical traits focusing on symptomatic participants, excluding data points where TFC score is 13 or ISS is 0 or 1. Results are adjusted for baseline value and age at motor symptom, with the outcome first RINTed by CAG, sex and education. Results are shown for POLD1 (top panel), PMS1 (middle), and FAN1 (bottom).

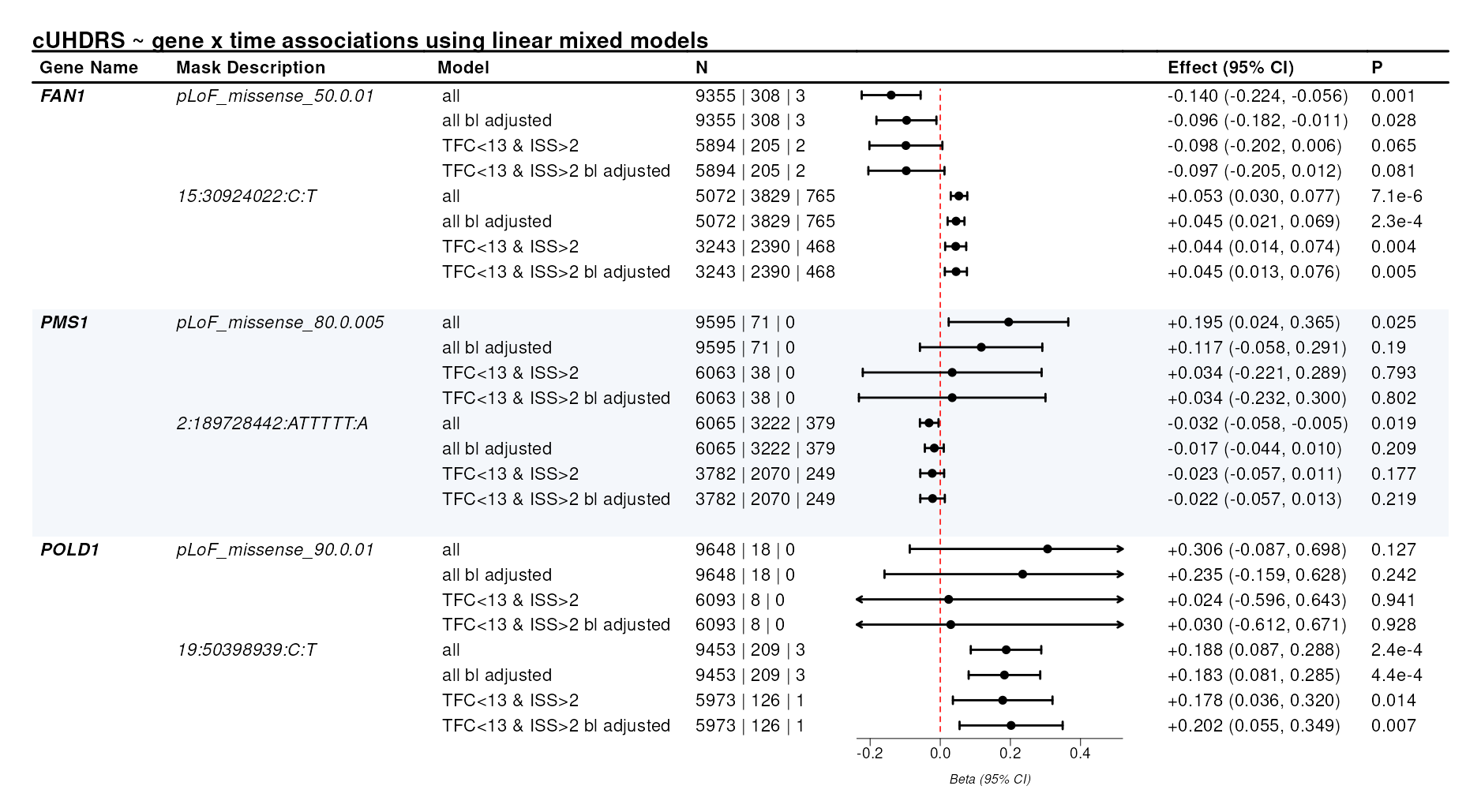

**Figure S3. cUHDRS progression using linear mixed models.** Forest plot of results for cUHDRS progression using linear mixed models, to assess additional modeling variations. Shown are summary statistics corresponding to the exposure (gene burden or variant) by time interaction in a model of longitudinal cUHDRS data including random intercepts and slopes by time for each participant. Each row for a given gene and mask description corresponds to a different (more stringent) modeling strategy.

To mimic the latter approach while enabling scalable GWAS, we also tested for the association between genotype and individual random slopes estimated for cUHDRS from a linear mixed model including age at first measure, time from baseline (in years) and number of pathogenic CAG repeats as fixed effect covariates. Random effects included random intercepts per participant and random slopes for the time variable. Individual random slopes were obtained from the models after adjusting and normalizing within CAG repeat groups prior to being used as quantitative traits for GWAS analyses. Although there were no genome-wide significant associations in this GWAS of cUHDRS progression (Figure S4A), effect sizes for the ten common variants identified in the GWAS of age at motor symptoms (Supplementary Figure 1) were correlated between the two analyses (Figure S4B). Consistent with this, a genetic risk score based on those ten variants had a significant association with the cUHDRS progression phenotype (Figure S4C).

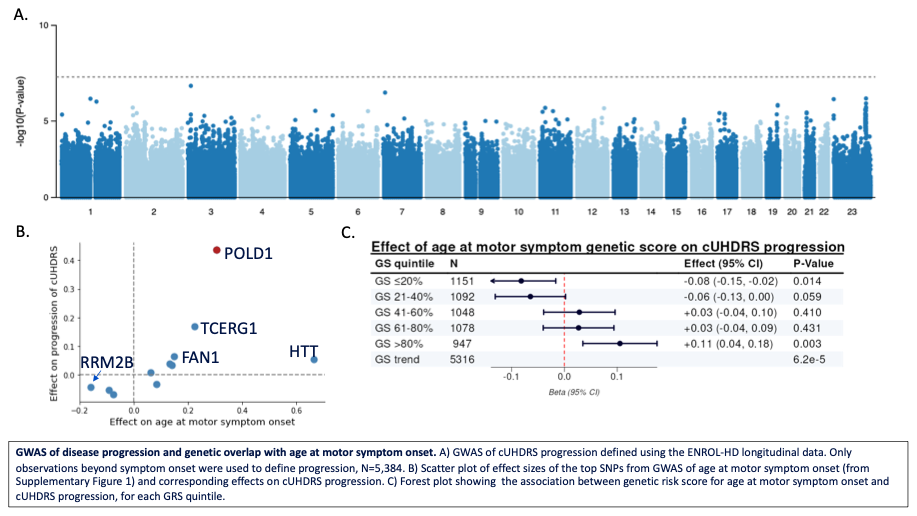

**Figure S4. Genetic associations with cUHDRS progression rate.** (A) GWAS of cUHDRS progression (N=5,384), defined using longitudinal data in the ENROL-HD cohort, using only observations recorded after symptoms onset. (B) Comparison of effect sizes estimated in the GWAS of motor symptom onset (x-axis) and the GWAS of cUHDRS progression (y-axis), considering the 10 lead variants identified in the former (see Supplementary Figure 1). (C) Association between cUHDRS progression and a genetic risk score calculated using the same 10 lead variants (weights from the GWAS of motor symptom onset). Linear regression was used to test for differences in cUHDRS progression between individuals in each genetic risk score quintile versus everyone else. The last row (“GS trend”) shows the significance of the overall association between cUHDRS progression and the genetic risk score as a continuous predictor.
