## Supplementary material for "Rare loss-of-function variants in *POLD1, PMS1* and *FAN1* modify age at onset of motor symptoms in Huntington’s disease": RGC banner

**RGC Management & Leadership Team**
Aris Baras, Gonçalo Abecasis, Adolfo Ferrando, Giovanni Coppola, Andrew Deubler, Luca A Lotta, John D Overton, Jeffrey G Reid, Alan Shuldiner, Katherine Siminovitch, Jason Portnoy, Marcus B Jones, Lyndon Mitnaul, Alison Fenney, Jonathan Marchini, Manuel Allen Revez Ferreira, Maya Ghoussaini, Mona Nafde, William Salerno, Cristen Willer, Lourdes Crane, Niek Verweij, Eric Jorgenson, and Joseph Pickrell.

**Sequencing & Lab Operations**
John D Overton, Christina Beechert, Erin Fuller, Laura M Cremona, Eugene Kalyuskin, Hang Du, Caitlin Forsythe, Zhenhua Gu, Kristy Guevara, Michael Lattari, Alexander Lopez, Kia Manoochehri, Prathyusha Challa, Manasi Pradhan, Raymond Reynoso, Ricardo Schiavo, Maria Sotiropoulos Padilla, Chenggu Wang, Sarah E Wolf, Hang Du, Kristy Guevara.

**Genome Informatics & Data Engineering**
Jeffrey G Reid, Mona Nafde, Manan Goyal, George Mitra, Sanjay Sreeram, Rouel Lanche, Vrushali Mahajan, Sai Lakshmi Vasireddy, Gisu Eom, Krishna Pawan Punuru, Sujit Gokhale, Shehroze Aamer, Pooja Mule, Mudasar Sarwar, Muhammad Aqeel, Xiaodong Bai, Lance Zhang, Sean O'Keeffe, Razvan Panea, Evan Edelstein, Devika Torvi, Ayesha Rasool, William Salerno, Evan K Maxwell, Boris Boutkov, Alexander Gorovits, Ju Guan, Alicia Hawes, Olga Krasheninina, Samantha Zarate, Adam J Mansfield, Lukas Habegger, Stephen Tahan, Naveen Karumuri.

**Analytical Genetics and Data Science**
Gonçalo Abecasis, Manuel Allen Revez Ferreira, Joshua Backman, Kathryn Burch, Adrian Campos, Liron Ganel, Sheila Gaynor, Benjamin Geraghty, Arkopravo Ghosh, Christopher Gillies, Lauren Gurski, Eric Jorgenson, Tyler Joseph, Michael Kessler, Jack Kosmicki, Adam Locke, Priyanka Nakka, Jonathan Marchini, Karl Landheer, Olivier Delaneau, Maya Ghoussaini, Anthony Marcketta, Joelle Mbatchou, Jonathan Ross, Carlo Sidore, Eli Stahl, Timothy Thornton, Rujin Wang, Kuan-Han Wu, Bin Ye, Blair Zhang, Andrey Ziyatdinov, Yuxin Zou, Jingning Zhang, Kyoko Watanabe, Mira Tang, Frank Wendt, Suganthi Balasubramanian, Suying Bao, Kathie Sun, Chuanyi Zhang, Sean Yu, Aaron Zhang, David Corrigan, Dhruv Shidhaye, Chen Wang, Keyrun Adhikari, Alexander Lachmann, Anna Alkelai, Mark Weiner, Julian Stamp, Joseph Pickrell, Ammar Tareen.

**Therapeutic Area Genetics**
Adolfo Ferrando, Giovanni Coppola, Luca A. Lotta, Alan Shuldiner, Katherine Siminovitch, Brian Hobbs, Jon Silver, William Palmer, Rita Guerreiro, Amit Joshi, Antoine Baldassari, Cristen Willer, Sarah Graham, Ernst Mayerhofer, Erola Pairo Castineira, Mary Haas, Niek Verweij, George Hindy, Jonas Bovijn, Tanima De, Luanluan Sun, Olukayode Sosina, Arthur Gilly, Peter Dornbos, Moeen Riaz, Manav Kapoor, Gannie Tzoneva, Veera Rajagopal, Sahar Gelfman, Vijay Kumar, Jacqueline Otto, Jose Bras, Silvia Alvarez, Jessie Brown, Hossein Khiabanian, Joana Revez, Kimberly Skead, Jae Soon Sul, Lei Chen, Sam Choi, Amy Damask, Nan Lin, Charles Paulding, Sameer Malhotra, Joseph Herman, Jacob McPadden, David Blair, Joshua Motelow, Julie Horowitz, Wenmin Zhang.

**Research Program Management & Strategic Initiatives**
Marcus B Jones, Michelle G LeBlanc, Nadia Rana, Jennifer Rico-Varela, Jaimee Hernandez, Larizbeth Romero, Ashley Paynter.

**Senior Partnerships & Business Operations**

Randi Schwartz, Lourdes Crane, Alison Fenney, Jody Hankins, Anna Han, Samuel Hart, Ryan Smith, Sarah Murphy.

**Business Operations & Administrative Coordinators**

Ann Perez-Beals, Gina Solari, Johannie Rivera-Picart, Michelle Pagan, Sunilbe Siceron.

Affiliations:

1. Regeneron Genetics Center, Tarrytown, NY, USA.
2. Contact information:
